# High risk of patient self-inflicted lung injury in COVID-19 with frequently encountered spontaneous breathing patterns: a computational modelling study

**DOI:** 10.1101/2021.03.17.21253788

**Authors:** Liam Weaver, Anup Das, Sina Saffaran, Nadir Yehya, Timothy E. Scott, Marc Chikhani, John G. Laffey, Jonathan G. Hardman, Luigi Camporota, Declan G. Bates

**Affiliations:** School of Engineering, University of Warwick, Coventry CV4 7AL, UK; Faculty of Engineering Science, University College London, London, WC1E 6BT, UK; Department of Anaesthesiology and Critical Care Medicine, Children’s Hospital of Philadelphia, University of Pennsylvania, Philadelphia, PA, USA; Academic Department of Military Anaesthesia and Critical Care, Royal Centre for Defence Medicine, ICT Centre, Birmingham B15 2SQ, UK; Anaesthesia & Critical Care, Division of Clinical Neuroscience, School of Medicine, University of Nottingham, Nottingham NG7 2UH, UK; Anaesthesia and Intensive Care Medicine, School of Medicine, NUI Galway, Ireland; Department of Critical Care, Guy’s and St Thomas’ NHS Foundation Trust, London, UK; Nottingham University Hospitals NHS Trust, Nottingham NG7 2UH, UK

**Author notes:** Equal contributions (Liam Weaver, Anup Das, Sina Saffaran).

**Keywords:** COVID-19, acute respiratory failure, hypoxaemia, patient self-inflicted lung injury, computational modelling

## Abstract

**Background:** There is on-going controversy regarding the potential for increased respiratory effort to generate patient self-inflicted lung injury (P-SILI) in spontaneously breathing patients with COVID-19 acute hypoxaemic respiratory failure. However, direct clinical evidence linking increased inspiratory effort to lung injury is scarce. We adapted a computational simulator of cardiopulmonary pathophysiology to quantify the mechanical forces that could lead to P-SILI at different levels of respiratory effort. In accordance with recent data, the simulator parameters were manually adjusted to generate a population of 10 patients that recapitulate clinical features exhibited by certain COVID-19 patients, i.e. severe hypoxaemia combined with relatively well-preserved lung mechanics, being treated with supplemental oxygen.

**Results:** Simulations were conducted at tidal volumes (VT) and respiratory rates (RR) of 7 ml/kg and 14 breaths/min (representing normal respiratory effort) and at VT/RR of 7/20, 7/30, 10/14, 10/20 and 10/30 ml/kg / breaths/min. While oxygenation improved with higher respiratory efforts, significant increases in multiple indicators of the potential for lung injury were observed at all higher VT/RR combinations tested. Pleural pressure swing increased from 12.0±0.3 cmH_2_O at baseline to 33.8±0.4 cmH_2_O at VT/RR of 7 ml/kg/30 breaths/min and to 46.2±0.5 cmH_2_O at 10 ml/kg/30 breaths/min. Transpulmonary pressure swing increased from 4.7±0.1 cmH_2_O at baseline to 17.9±0.3 cmH_2_O at VT/RR of 7 ml/kg/30 breaths/min and to 24.2±0.3 cmH_2_O at 10 ml/kg/30 breaths/min. Total lung strain increased from 0.29±0.006 at baseline to 0.65±0.016 at 10 ml/kg/30 breaths/min. Mechanical power increased from 1.6±0.1 J/min at baseline to 12.9±0.2 J/min at VT/RR of 7 ml/kg/30 breaths/min, and to 24.9±0.3 J/min at 10 ml/kg/30 breaths/min. Driving pressure increased from 7.7±0.2 cmH_2_O at baseline to 19.6±0.2 at VT/RR of 7 ml/kg/30 breaths/min, and to 26.9±0.3 cmH_2_O at 10 ml/kg/30 breaths/min.

**Conclusions:** Our results suggest that the forces generated by increased inspiratory effort commonly seen in COVID-19 acute hypoxaemic respiratory failure are comparable with those that have been associated with ventilator-induced lung injury during mechanical ventilation. Respiratory efforts in these patients should be carefully monitored and controlled to minimise the risk of lung injury.

## Introduction

On admission, some patients with COVID-19 acute hypoxaemic respiratory failure (AHRF) exhibit profound hypoxaemia, combined with relatively preserved lung compliance and lung gas volume on CT chest imaging, and substantial increases in respiratory effort - tidal volumes (VT) of 15-20 ml/kg (1) and respiratory rates (RR) of 34 breaths/min (2) have been reported. As noted in (3), young, otherwise healthy adults can sustain tidal volumes of 20 ml/kg at a respiratory rate of 45 breaths/min almost indefinitely (4,5). There is significant debate regarding whether sustained high respiratory effort in these patients could risk causing further damage to the lungs through patient self-inflicted lung injury (P-SILI) (6–11).

Direct evidence for the existence of P-SILI in the context of purely spontaneous breathing is largely based on an animal study (12), although two studies in asthmatic children suggested that increased breathing effort could promote negative pressure pulmonary oedema (13, 14). A number of studies have also established the potential for injurious effects due to spontaneous breathing during mechanical ventilation in acute respiratory failure (15,16). In a recent study of inspiratory effort in non-invasive ventilation, reductions in the oesophageal pressure swings (pleural pressure) of 10 cm H_2_O or more after 2 hours of treatment was strongly associated with avoidance of intubation and represented the most accurate predictor of treatment success (17–19). In the context of COVID-19, a recent study has asserted an association between increased respiratory effort and worsening of respiratory function during attempts to wean patients from mechanical ventilation, although without definitively establishing a delineation between cause and effect (20,10,11). Two recent case reports also noted the existence of spontaneous pneumothorax and pneumomediastinum in COVID-19 patients, suggesting the generation of injurious transpulmonary pressures (21,22).

To obtain some additional evidence, we hypothesised that a computational model of COVID-19 pathophysiology could be used to investigate the effects of increased respiratory effort on parameters that have been associated with lung injury, namely tidal swings in pleural and transpulmonary pressure, and maximum values of mechanical power (23), driving pressure and lung strain. The aim of the study was to quantify the levels of these indicators of lung injury that are generated in our model by breathing patterns that are frequently encountered in COVID-19 patients.

## Methods

### Core model

The core model used in this study is a multi-compartmental computational simulator that has been previously used to simulate mechanically ventilated patients with various pulmonary disease states (24–31), including COVID-19 ARDS (32). The simulator offers several advantages, including the capability to define a large number of alveolar compartments (each with its own individual mechanical characteristics), with configurable alveolar collapse, alveolar stiffening, disruption of alveolar gas-exchange, pulmonary vasoconstriction and vasodilation, and airway obstruction. As a result, several defining clinical features of acute lung injury can be represented in the model, including varying degrees of ventilation perfusion mismatch, physiological shunt and deadspace, alveolar gas trapping with intrinsic positive end-expiratory pressure (PEEP), collapse-reopening of alveoli etc. A detailed description of the physiological principles and mathematical equations underlying the core computational model implemented in the simulator is provided in the supplementary file. A list of key parameters in the model is given in Table S1.1 of the SM.

### Adaptation to COVID-19 pathophysiology

For the current study, the model was configured to represent a patient of 70 kg ideal body weight with COVID-19 acute hypoxaemic respiratory failure, as follows.

Based on recent data [1,33–38] suggesting that some COVID-19 patients have relatively well preserved lung gas volume and compliance, the model was set to have 8% of its alveolar compartments collapsed, i.e. non-aerated. To simulate the hyperperfusion of gasless tissue reported in [1,39,40], we implemented vasodilation in the collapsed units by decreasing their vascular resistance by 80%. HPV is normally incorporated in our simulator via a mathematical function, to simulate the hypothesised disruption of HPV in COVID-19 we disabled this function in our model. We incorporated disruption of alveolar gas-exchange due to the effects of pneumonitis into the model by blocking alveolar-capillary gas equilibration in 20% of the alveolar compartments. As thrombotic complications have been reported to be a characteristic feature of COVID-19 [41,42], we also modeled the presence of microthrombi by increasing vascular resistance by a factor of 5 in 10% of the remaining compartments.

Implementing the above pathophysiological mechanisms in our model produced levels of shunt (49.5%) and deadspace (188.5 ml) leading to severe hypoxemia (SaO_2_ 84%, FiO_2_ of 100%) consistent with recent clinical data on COVID-19 ARDS, while still maintaining relatively well preserved levels of respiratory system compliance (63 ml/cm H_2_O) – see SM Table S5.1. To ensure that our results are not dependent on this particular model parameterization, we then created a “population” of 10 patients by varying the number of compartments affected by the various pathophysiological mechanisms around this nominal parameterisation, producing a range of hypoxemia/compliance levels that are still consistent with reported data; see SM, Table S5.1.

### Modelling spontaneous breathing

For the current investigation, the core model was adapted to represent spontaneously breathing rather than mechanically ventilated patients. Spontaneous breathing is simulated by incorporating the variable *P*_*INSP*_, which represents the lumped effect of chest wall, muscle and pleura on the lung. During a single respiratory cycle, *P*_*INSP*_ at time t_k_ is calculated by adapting a model developed in (43,44) based on breathing profiles of 12 patients:

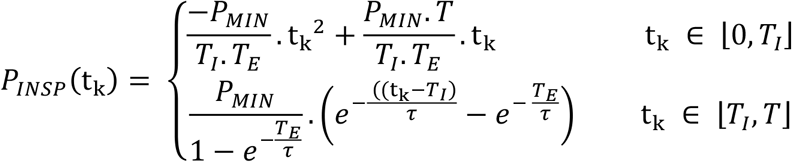

The function consists of a parabolic profile during the inspiration phase of the respiratory cycle, representing the progressive increase in pressure exerted by the respiratory muscles. This is followed by an exponential profile during the expiration phase of the respiratory cycle, characterizing the passive relaxation of the muscles (a valid assumption up to a minute ventilation of 40 l/min, (45)). *P*_*INSP*_ decreases from zero to its minimum end-inspiratory value (*P*_*MIN*_) (i.e., maximum effort) during inspiration and returns to zero at end of expiration. *T* is calculated from the set respiratory rate, RR, (*T* = 60/RR). *T*_I_ and *T*_*E*_ are the duration of inspiration and expiration, such that (*T* = *T*_*I*_ + *T*_*E*_). *T*_*I*_ is calculated from (*T*_*I*_ = *T* * *DC*) where *DC* is the duty cycle, set to 0.33. *τ* is the time constant of the expiratory profile and is set to *T*_*E*_/RR seconds.

### Calculating indices of lung injury

Pressures inside the lung are calculated in the model in an iterative fashion, as follows:

Step 1. Calculate *p*_*i*_, the pressures inside each individual alveolar unit using the equation:

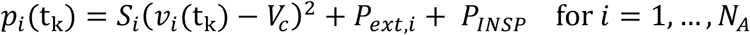

The parameter *S*_*i*_ reflects the ‘stiffness’ of the compartment, *P*_*ext,i*_ represents the effective net pressure acting on the alveolar compartments due to assorted localised mechanisms (e.g. oedema formation), and *V*_*c*_ is defined as the volume at which the alveolar compartment is considered to be ‘empty’, to avoid divide by 0 errors.

Step 2. Calculate *P*_*L*_ using the values of alveolar pressures, *p*_*i*_, and airway resistances *R*_*Bi*_, as follows: assuming zero net flow between mouth and lungs, calculate the separate alveolar flows (*p*_*i*_)/ *R*_*Bi*_, add these together to get total flow, calculate total airway resistance, and divide total flow by total airway resistance to get global lung pressure. The values of *R*_*Bi*_ are fixed and set to represent different pathophysiological mechanisms in the model.

Step 3. Calculate the respiratory system compliance *C*_*rs*_ from

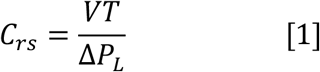

where *VT* is the tidal volume, and Δ*P*_*L*_ is the difference between the minimum lung pressure and end expiratory lung pressure. Note that Equation 1 gives us *C*_*rs*_, rather than *C*_*L*_, because we are applying *P*_*ext*_ and *P*_*INSP*_ to the alveoli directly (an anatomical modeling simplification but of no consequence mathematically).

Step 4. Calculate the dynamic lung compliance *C*_*L*_

Since *EL*_*rs*_ = *EL*_*L*_ + *EL*_*cw*_, we have that 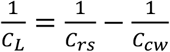

where *C*_*cw*_ is the chest wall compliance (set to 144 mL/cmH_2_O based on data from COVID-19 patients in (46)) and *C*_*rs*_ has been found from Eq. 1.

Step 5. Calculate transpulmonary pressure *P*_*tp*_ (equivalent to lung stress (47))

Since 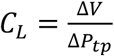, then

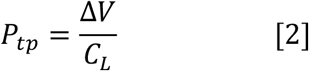

if Δ*P*_*tp*_= *P*_*tp*_ − 0, and Δ*V* is the difference between the current lung volume and the unstressed volume of the lung (the residual volume, set to 1.2L).

Step 6. Calculate pleural pressure *P*_*pl*_ from: *P*_*pl*_ = *P*_*L*_ – *P*_*tp*_

Step 7. Calculate the mechanical power applied to the lungs as the energy per breath (defined as the area of the triangle enclosed by the inspiratory limb of the Δ*P*_*tp*_ pressure-volume curve and the change in volume, i.e. 0.5×*VT*×Δ*P*_*tp*_, in Joules) multiplied by RR, as described in (48).

Step 8. Calculate driving pressure Δ*P* as Δ*P* = *VT*×*EL*_*RS*_ = Δ*P*_*L*_ (from Equation 1).

Step 9. Calculate the total lung strain as the sum of dynamic and static strain (49), where

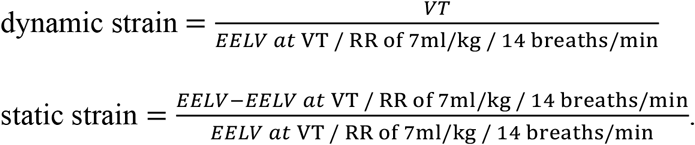

To observe the various pulmonary effects of interest, the following values were also computed and recorded: arterial oxygen partial pressure (PaO_2_), arterial carbon dioxide partial pressure (PaCO_2_), physiological shunt, calculated using the classical shunt equation, based on the calculated values of arterial, pulmonary end-capillary and mixed venous O_2_ content. Physiological deadspace (and deadspace fraction; VD/VT) were calculated from PaCO_2,_ mixed expired CO_2_ pressure (*PĒCO*_2_) and the exhaled tidal volume.

All model simulations were run for 30 minutes, with the reported data averaged over the final 1 minute, and conducted using Matlab version R2019b.v9 (MathWorks Inc., Natick, MA, USA).

## Results

The simulated patient population replicates levels of hypoxaemia that have frequently been reported in spontaneously breathing COVID-19 patients. SaO_2_, PaO_2_ and PaCO_2_ on 100% oxygen at baseline were 83.8±5.1%, 52.6±7.1 mmHg, and 51.8±1.5 mmHg, respectively (Table 1). Oxygenation improved at higher respiratory efforts (SaO_2_ of 97.6±0.8 at VT / RR of 10 ml/kg / 30 breath/min) and PaCO_2_ decreased. Values of respiratory system compliance and end-expiratory lung volume (EELV) at baseline were 63±1.4 ml/cmH_2_O and 1678.9±35.3 ml, showing good agreement with the data for a cohort of COVID-19 patients reported in a recent study (38). Compliance reduced significantly at higher respiratory rates (to 25±0.3 ml/cmH_2_O at VT / RR of 7 ml/kg / 30 breath/min). Physiological shunt and deadspace were decreased and increased, respectively, at higher respiratory effort.

**Table 1:**
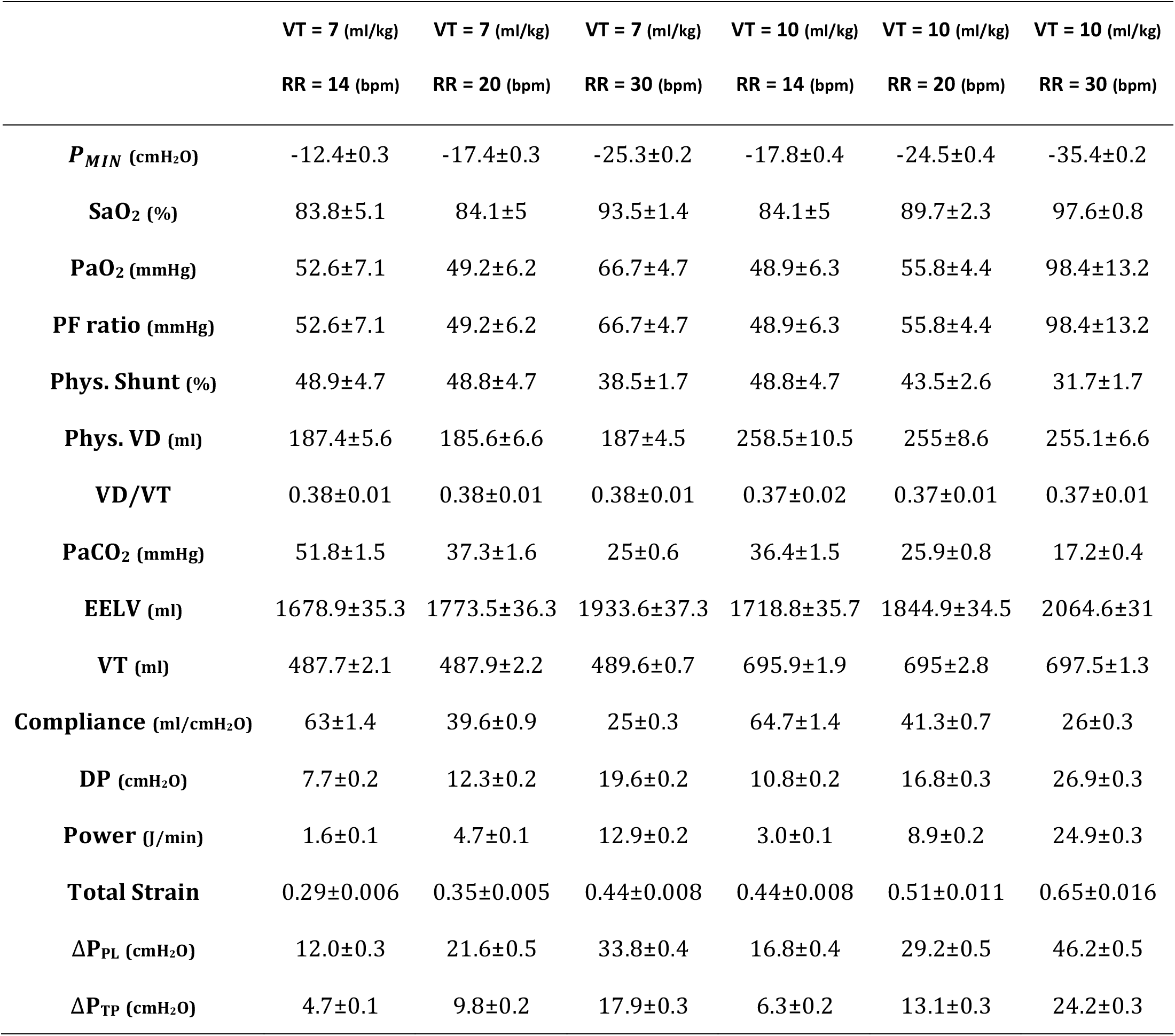
Oxygenation, lung mechanics, maximum pressure swings, and other injury indicators generated in our COVID-19 model for different levels of respiratory effort on 100% oxygen, data presents mean ± sd across the population of 10 patients.

Inspiratory pressures required to generate the different increased breathing patterns (maximum *P*_*MIN*_ of 35.4±0.2 cmH_2_O at VT / RR of 10 ml/kg / 30 breath/min) were well within the limits specified for a 70kg adult male (*P*_*MIN*_ of 78.5 cmH_2_O at age 60, and 108.9 cmH_2_O at age 40, (50)).

As shown in Table 1 and Figure 1, significant increases in multiple indicators of the potential for lung injury were observed at all higher VT / RR combinations tested. Pleural pressure swing increased from 12.0±0.3 cmH_2_O at baseline to 33.8±0.4 cmH_2_O at VT/RR of 7 ml/kg / 30 breaths/min and to 46.2±0.5 cmH_2_O at 10 ml/kg / 30 breaths/min. Transpulmonary pressure swing increased from 4.7±0.1 cmH_2_O at baseline to 17.9±0.3 cmH_2_O at VT/RR of 7 ml/kg / 30 breaths/min and to 24.2±0.3 cmH_2_O at 10 ml/kg / 30 breaths/min. Total lung strain increased from 0.29±0.006 at baseline to 0.65±0.016 at 10 ml/kg / 30 breaths/min. Mechanical power increased from 1.6±0.1 J/min at baseline to 12.9±0.2 J/min at VT/RR of 7 ml/kg / 30 breaths/min, and to 24.9±0.3 J/min at 10 ml/kg / 30 breaths/min. Driving pressure increased from 7.7±0.2 cmH_2_O at baseline to 19.6±0.2 at VT/RR of 7 ml/kg / 30 breaths/min, and 26.9±0.3 cmH_2_O at 10 ml/kg / 30 breaths/min.

**Figure 1:**
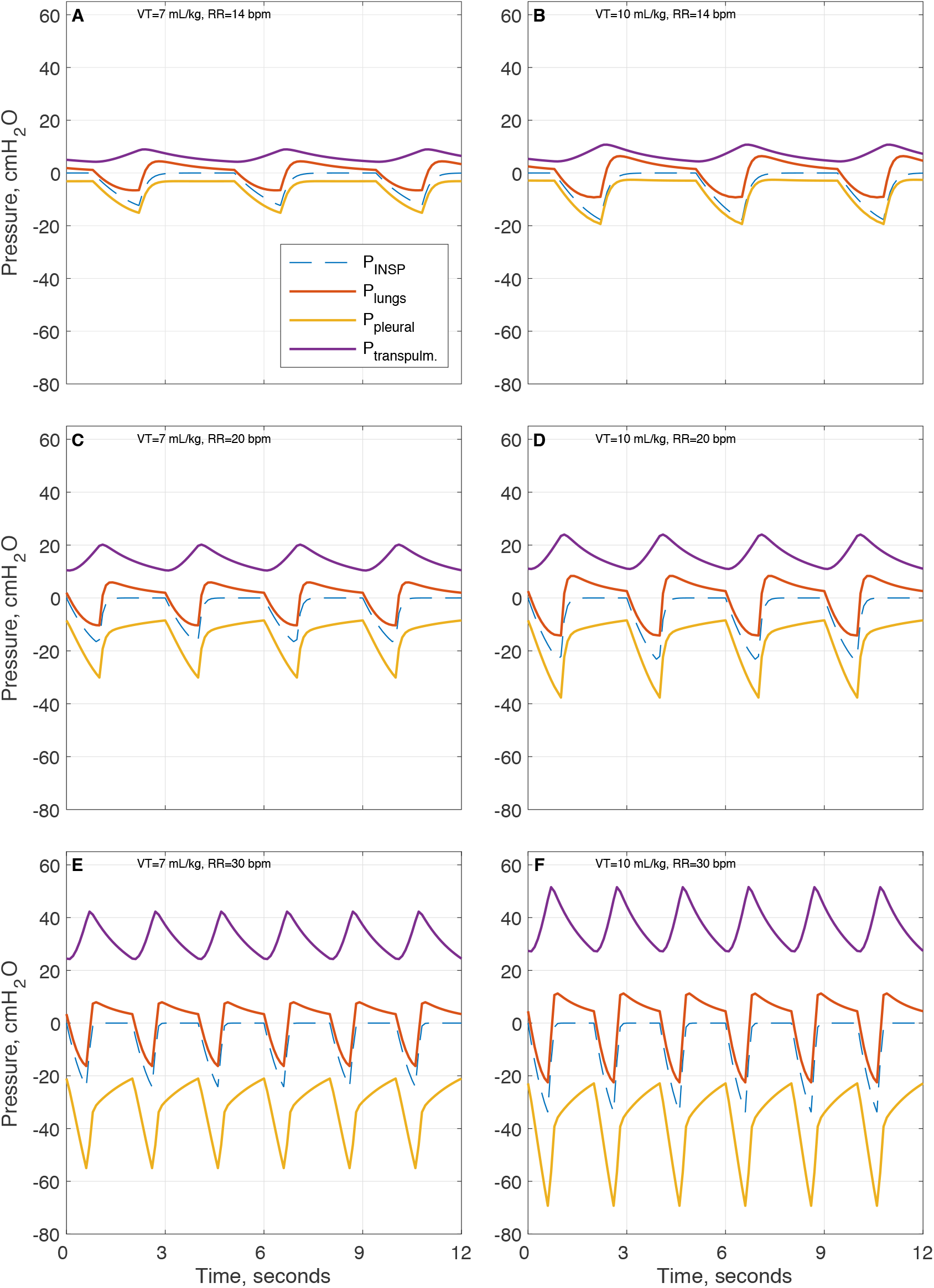
Inspiratory, pleural, transpulmonary and alveolar pressure swings generated by normal and increased respiratory effort in our COVID-19 model (patient 1 – see SM Table S5.1), (A) VT = 7 ml/kg, RR = 14 b/min, (B) VT = 10 ml/kg, RR = 14 b/min, (C) VT = 7 ml/kg, RR = 20 b/min, (D) VT = 10 ml/kg, RR = 20 b/min, (E) VT = 7 ml/kg, RR = 30 b/min, (F) VT = 10 ml/kg, RR = 30 b/min.

The effect of increased respiratory effort on the distribution of maximum compartmental volumes in the model is shown in Fig. S4.1 and Fig. S4.2 in the supplementary file. As shown, at the higher values of tidal volume and respiratory rate, the proportion of model compartments experiencing larger maximum volumes is significantly increased.

## Discussion

During mechanical ventilation, the power required to inflate the lungs is provided by an external source of energy, whereas during spontaneous unassisted breathing it is provided by the respiratory muscles. However, as pointed out in (51), lung injury (in the sense of mechanical lesions in the interstitial space due to microfractures of the extracellular matrix or the capillary walls) arises from the mechanical energy applied to the lungs, which generates the relevant pressures. There is therefore no reason to believe that the extent of injury will be significantly different whether excessive pressures are generated by respiratory muscles in spontaneous breathing or by a mechanical ventilator. Similarly to the case of VILI, there is also no reason to expect that, in the case of injured lungs, forces generated by respiratory muscles could not lead to injurious effects on a regional level due to lung heterogeneity. This may be a particular concern in the case of COVID-19 AHRF, since current understanding of the underlying pathophysiology points to a strongly heterogeneous lung profile incorporating alveolar collapse, oedema, and significant vascular derangement.

As well as increases in transpulmonary pressure swings, we also observed large increases in pleural pressure swings at higher respiratory effort, up to a maximum of 46.2±0.5 cmH_2_O. As discussed in (16), negative alveolar pressures created by large changes in pleural pressure, and therefore positive changes in transvascular pressure, favour lung oedema, a mechanism that is amplified with increased vascular permeability, (52, 53). Given that negative pressures from diaphragm contraction are not distributed uniformly, there is also the potential to cause pendelluft gas movement due to localised changes in pleural pressures in dependent regions, (54). Finally, it is important to recognise that when dynamic systems (from aircraft engines to human lungs) are subjected to repeated cycles of excessive stresses and strains, their deterioration over time is often not linear; rather, damage can accumulate “silently” before eventually manifesting and spreading rapidly (55).

In light of the above considerations, it is difficult to see how, for a respiratory effort that a patient (and their treating clinician) might consider tolerable, the levels of transpulmonary/pleural pressure swings, driving pressure and mechanical power produced in our model (Table 1) could be regarded as safe. Certainly, it is unlikely that any mechanical ventilation strategy that produced similar values would be considered “protective” according to current standard guidelines. Our results also highlight that improvements in PaO_2_/FiO_2_ ratio associated with larger tidal volumes and respiratory rates should be interpreted in the context of the associated increased respiratory effort, and are not necessarily expressions of improved lung condition. Indeed, the improvement in PaO_2_/FiO_2_ ratio we observed at higher respiratory effort was coupled with reduced lung compliance and the development of levels of mechanical power associated with worse short (56, 57) and long term (58) survival.

Our study has a number of limitations. The results are based on computational modelling of mechanisms that have been proposed to underlie COVID-19 pathophysiology, rather than on models matched to individual data from patients with COVID-19. Accordingly, many model parameters were manually adjusted to give overall outputs that are similar to reported data on COVID-19 patients, rather than being fit to data that explicitly defines the parameters. The model also neglects some physiological realities (namely interdependence of alveoli, non-uniformity of diaphragm contraction, and gravitational effects), however, it seems reasonable to expect that inclusion of these effects would act to produce even higher localised values of the reported lung injury indices in certain lung regions. Due to a lack of data we have also not modelled the potential effects of variations in the ratio of inspiratory/expiratory time at higher respiratory rates.

Our results are limited to the case of purely spontaneous breathing with oxygen support – consideration of the effects of high respiratory effort during positive pressure non-invasive ventilation (CPAP, BIPAP, etc) is an important open question, but requires further development of the model and will be the subject of future studies.

Investigating the issues raised here in clinical trials is likely to prove challenging both from an ethical and practical point of view, and suitable animal models of COVID-19 pathophysiology with which to study these questions are yet to emerge (59). In these circumstances, we hope that insights from detailed computational models that recapitulate patient breathing patterns and lung mechanics can provide useful evidence with which to inform current and future debates (60).

## Conclusions

Our results indicate that transpulmonary and pleural pressure swings, and levels of driving pressure, lung strain and mechanical power that have been associated with VILI during mechanical ventilation can develop in spontaneously breathing patients with COVID-19 acute respiratory failure, at levels of respiratory effort that are being frequently encountered by clinicians. Respiratory efforts in these patients should be carefully monitored and controlled to minimise the risk of lung injury.

## Data Availability

Requests for data and further details could be forwarded to

## Supplementary Materials

AIC_Supplementary.pdf – Full description of computational simulator, additional results and figures.

## Availability of data and materials

All data for the study is contained in the paper and the SM. Requests to be forwarded to Prof. Declan G Bates.

## Ethics approval and consent to participate

Not applicable

## Consent for publication

Not applicable

## Authors’ contributions

DGB designed the study. AD, LW and SS performed the modelling and carried out simulations. All authors contributed to the modelling, analysed the data, and contributed to writing the paper.

## Acknowledgements

Not applicable.

## Authors’ information (optional)

Not applicable.

