## Supplemental File for "High risk of patient self-inflicted lung injury in COVID-19 with frequently encountered spontaneous breathing patterns: a computational modelling study"

### Additional File

The online data supplement for this paper contains additional material that could not be included in the main text due to space limitations. The following sections describe in detail the simulation model employed in the paper.

### Section 1: ICSM Pulmonary Model

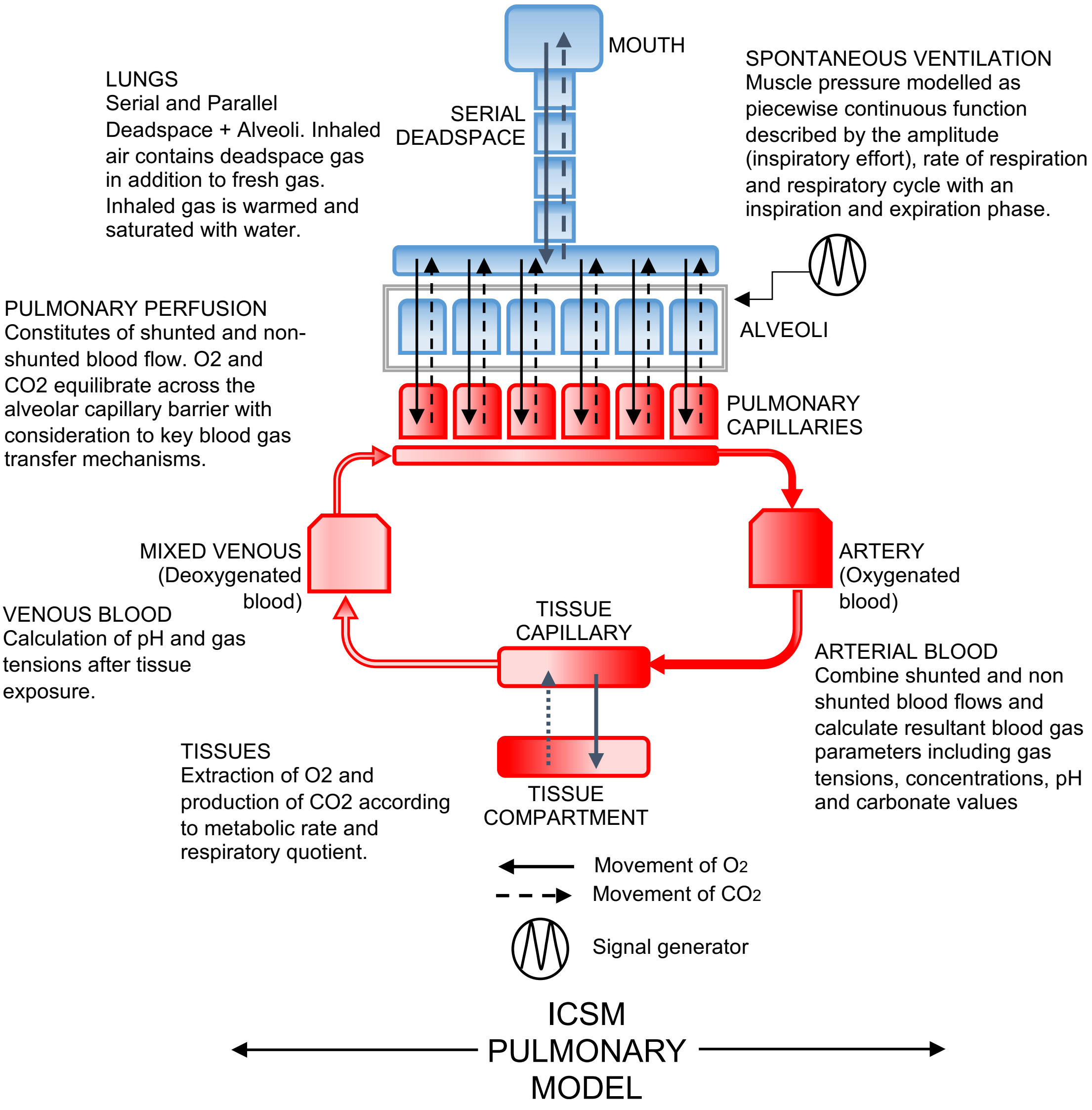

**Figure S1.1 Diagrammatic representation of the pulmonary model**

The model employed in this paper has been developed over the past several years and has been applied and validated on a number of different studies (1-8). The model is organised as a system of several components (see Fig S1.1), each component representing different sections of pulmonary dynamics and blood gas transport, e.g. the transport of air in the mouth, the tidal flow in the airways, the gas exchange in the alveolar compartments and their corresponding capillary compartment, the flow of blood in the arteries, the veins, the cardiovascular compartment, and the gas exchange process in the peripheral tissue compartments. Each component is described as several mass conserving functions and solved as algebraic equations, obtained or approximated from the published literature, experimental data and clinical observations. These equations are solved in series in an iterative manner, so that solving one equation at current time instant ($t_k$) determines the values of the independent variables in the next equation. At the end of the iteration, the results of the solution of the final equations determine the independent variables of the first equation for the next iteration.

The iterative process continues for a predetermined time, *T*, representing the total simulation time, with each iteration representing a 'time slice' $t$ of real physiological time (set to 10 ms). At the first iteration$(t_k, k = 0)$, an initial set of independent variables are chosen based on values selected by the user. The user can alter these initial variables to investigate the response of the model or to simulate different pathophysiological conditions. Subsequent iterations $(t_k = t_{k-1} + t)$ update the model parameters based on the equations below.

The pulmonary model consists of the mechanical ventilation equipment, anatomical and alveolar dead space, anatomical and alveolar shunts, ventilated alveolar compartments and corresponding perfused capillary compartments. The pressure differential created by the mechanical ventilator drives the flow of gas through the system. The series dead space (SD) is located between the mouth and the alveolar compartments and consists of the trachea, bronchi and the bronchioles where no gas exchange occurs. Inhaled gases pass through the SD during inspiration and alveolar gases pass through the SD during expiration. In the model, an SD of volume 60ml is split into 50 stacked layers of equal volumes ($N_{SD}$ = 50). No mixing between the compartments of the SD is assumed.

Any residual alveolar air in the SD at the end of expiration is re-inhaled as inspiration is initiated. This residual air is composed of gases exhaled from both perfused alveolar compartments (normal perfusion) and the parallel dead space (PD) (alveolar compartments with limited perfusion). Therefore, the size of dead space (SD and PD) can have a significant effect on the gas composition of the alveolar compartments.

The inhaled air is initially assumed to consist of five gases: oxygen ($O_2$), nitrogen ($N_2$), carbon dioxide ($CO_2$), water vapor ($H_2O$) and a 5th gas (α) used to model additives such as helium or other anaesthetic gases. During an iteration of the model, the flow (*f*) of air to or from an alveolar compartment *i* at time $t_k$is determined by the following equation:

$$f_i(t_k) = \frac{(p_v(t_k) - p_i(t_k))}{(R_u + R_{A,i})} \qquad \text{for } i = 1, \dots, N_A \qquad (1)$$

where $p_v(t_k)$ is the pressure supplied by the mechanical ventilator at $(t_k)$, $p_i(t_k)$ is the pressure in the alveolar compartment $i$ at $(t_k)$, $R_u$ is the constant upper airway resistance and $R_{A,i}$ is the bronchial inlet resistances of the alveolar compartment $i$. $N_A$ is the total number of alveolar compartments (for the results in this paper, $N_A$ = 100). The total flow of air entering the SD at time $t_k$ is calculated by

$$f_{SD}(t_k) = \sum_{i=1}^{N_A} f_i(t_k) \qquad (2)$$

During the inhaling phase, $f_{SD} \geq 0$, while in the exhaling phase $f_{SD} < 0$. During gas movement in the SD, the fractions of gases in the layer $l$ of the SD, $\mathrm{F}_{l,}$ $(l = 1, \ldots, N_{SD})$ is updated based on the composition of the total flow, $f_{SD}$ , and the current composition of $\mathrm{F}_{l,}$. If $f_{SD}$ ≥ 0, then air starts filling from the top layer ($l$ = 1) to the bottom layer ($l = N_{SD}$); and vice versa for $f_{SD}$< 0.

The volume of gas x, in the $i^{th}$ alveolar compartment ($\mathrm{v}_{i,\,\mathrm{x}}$), is given by:

$$v_{i,\mathrm{x}}(\mathrm{t_k}) = \begin{cases} v_{i,\mathrm{x}}(\mathrm{t_{k-1}}) - f_i(\mathrm{t_k}) \cdot \frac{v_{i,\mathrm{x}}(\mathrm{t_{k-1}})}{v_i(\mathrm{t_k})} & \mathit{Exhaling} \\ v_{i,\mathrm{x}}(\mathrm{t_{k-1}}) + f_i(\mathrm{t_k}) \cdot \mathrm{F}_{N_{SD}}(\mathrm{t_k}) & \mathit{Inhaling} \end{cases} \quad \text{for } i = 1, \ldots, N_A \qquad (3)$$

In (3), x is any of the five gases ($O_2$, $N_2$, $CO_2$, $H_2O$ or α). The total volume of the $i^{th}$ alveolar compartment, $v_i$ is the sum of the volume of the five gases in the compartment.

$$v_i(\mathrm{t_k}) = v_{i,\mathrm{O2}}(\mathrm{t_k}) + v_{i,\mathrm{N2}}(\mathrm{t_k}) + v_{i,\mathrm{CO2}}(\mathrm{t_k}) + v_{i,\mathrm{H2O}}(\mathrm{t_k}) + v_{i,\alpha}(\mathrm{t_k}) \qquad (4)$$

For the alveolar compartments, the tension at the centre of the alveolus and at the alveolar capillary border is assumed to be equal. The respiratory system has an intrinsic response to low oxygen levels in blood which is to restrict the blood flow in the pulmonary blood vessels, known as hypoxic pulmonary vasoconstriction (HPV). This is modelled as a simple function, resembling the stimulus response curve suggested by Marshall (9), and is incorporated into the simulator to gradually constrict the blood vessels as a response to low alveolar oxygen tension. The atmospheric pressure is fixed at 101.3 kPa and the body temperature is fixed at 37.2°C.

At each $\mathrm{t_k}$, equilibration between the alveolar compartment and the corresponding capillary compartment is achieved iteratively by moving small volumes of each gas between the compartments until the partial pressures of these gases differ by <1% across the alveolar-capillary boundary. The process includes the nonlinear movement of $O_2$ and $CO_2$ across the alveolar capillary membrane during equilibration.

In blood, the total $O_2$ content ($C_{O2}$) is carried in two forms, as a solution and as oxyhaemoglobin (saturated haemoglobin):

$$\mathrm{C_{O2}}(\mathrm{t_k}) = \mathrm{S_{O2}}(\mathrm{t_{k-1}}) \cdot Huf \cdot \mathrm{Hb} + \mathrm{P_{O2}}(\mathrm{t_{k-1}}) \cdot \mathrm{O}_{2sol} \qquad (5)$$

In this equation, $S_{O2}$ is the haemoglobin saturation, *Huf* is the Hufner constant, Hb is the haemoglobin content and $\mathrm{O}_{2sol}$ is the $O_2$ solubility constant. The following pressure-saturation relation, as suggested by (10) to describe the $O_2$ dissociation curve, is used in this model:

$$\mathrm{S_{O2}}(\mathrm{t_k}) = \left(\left(\left(\mathrm{P_{O2}^3}(\mathrm{t_{k-1}}) + 150 \cdot \mathrm{P_{O2}}(\mathrm{t_{k-1}})\right)^{-1} \times 23400\right) + 1\right)^{-1} \qquad (6)$$

$S_{O2}$ is the saturation of the haemoglobin in blood and $P_{O2}$is the partial pressure of oxygen in the blood. As suggested by (11), $P_{O2}$ has been determined with appropriate correction factors in base excess BE, temperature T and pH (7.5005168 = pressure conversion factor from kPa to mmHg):

$$P_{O2}(t_k) = 7.5006168 \cdot P_{O2}(t_{k-1}) \cdot 10^{[0.48(pH(t_{k-1})-7.4)-0.024(T-37)-0.0013 \cdot BE]} \qquad (7)$$

The $CO_2$ content of the blood ($C_{CO2}$) is deduced from the plasma $CO_2$ content ($C_{CO2plasma}$) (12) by the following equation:

$$C_{CO2}(t_k) = C_{CO2plasma}(t_{k-1}) \cdot \left[1 - \frac{0.0289 \cdot Hb}{(3.352 - 0.456 . S_{O2}(t_k)) \cdot (8.142 - pH(t_{k-1}))}\right] \qquad (8)$$

where $S_{O2}$ is the $O_2$ saturation, Hb is the haemoglobin concentration and pH is the blood pH level. The coefficients were determined as a standardized solution to the McHardy version of Visser's equation (13), by iteratively finding the best fit values to a given set of clinical data. The value of $C_{CO2plasma}$ is deduced using the Henderson-Hasselbach logarithmic equation for plasma $C_{CO2}$ (14):

$$C_{CO2plasma}(t_k) = 2.226 \cdot s_{CO2} \cdot P_{CO2}(t_{k-1}) \left(1 + 10^{(pH(t_{k-1}) - pK')}\right) \qquad (9)$$

where $s_{CO2}$ is the plasma $CO_2$ solubility coefficient and pK' is the apparent pK (acid dissociation constant of the $CO_2$ bicarbonate relationship). $P_{CO2}$ is the partial pressure of $CO_2$ in plasma and '2.226' refers to the conversion factor from millimoles per liter to ml/100ml. (14) gives the equations for $s_{CO2}$ and pK' as:

$$s_{CO2} = 0.0307 + 0.0057 \cdot (37 - T) + 0.00002 \cdot (37 - T)^2 \qquad (10)$$

$$pK' = 6.086 + 0.042 \cdot (7.4 - pH(t_{k-1})) + (38 - T) \cdot \left(0.00472 + \left(0.00139 - (7.4 - pH(t_{k-1}))\right)\right) \qquad (11)$$

$P_{CO2}(t_k)$ is determined by incorporating the standard Henry's law and the $s_{CO2}$ (the $CO_2$ solubility coefficient above). For pH calculation, the Henderson Hasselbach and the Van Slyke equation (15) are combined. Below is the derivation of the relevant equation. The Henderson-Hasselbach equation (governed by the mass action equation (acid dissociation)) states that:

$$pH = pK + \log\left(\frac{bicarbonateconcentration}{carbonicacidconcentration}\right) \qquad (12)$$

Substituting pK=6.1 (under normal conditions) and the denominator $(0.225 \cdot P_{CO2})$ (acid concentration being a function of $CO_2$ solubility constant 0.225 and $P_{CO2}$ (in kPa)) gives:

$$pH(t_k) = 6.1 + \log\left(\frac{HCO_3(t_{k-1})}{0.225 \cdot P_{CO2(t_k)}}\right) \qquad (13)$$

For a given pH, base excess (BE), and haemoglobin content (Hb), $HCO_3$ is calculated using the Van-Slyke equation, as given by(15):

$$HCO_3(t_k) = \left((2.3 \times Hb + 7.7) \times (pH(t_k) - 7.4)\right) + \frac{BE}{(1 - 0.023 \times Hb)} + 24.4 \qquad (14)$$

The capillary blood is mixed with arterial blood using the equation below which considers the anatomical shunt ($Sh$) with the venous blood content of gas x ($C_{v,x}$), the non-shunted blood content from the pulmonary capillaries ($C_{cap,x}$), arterial blood content ($C_{a,x}$), the arterial volume ($v_a$) and the cardiac output (CO).

$$C_{a,x}(t_k) = \frac{CO(t_k) \cdot (Sh \cdot C_{v,x}(t_k) + (1-Sh) \cdot C_{cap,x}(t_k)) + C_{a,x}(t_k) \cdot (v_a(t_k) - CO(t_k))}{v_a(t_k)} \quad (15)$$

The peripheral tissue model consists of a single tissue compartment, acting between the peripheral capillary and the active tissue (undergoing respiration to produce energy). The consumed $O_2$ ($V_{O2}$) is removed and the produced $CO_2$ ($V_{CO2}$) is added to this tissue compartment. As per the alveolar equilibration, peripheral capillary gas partial pressures reach equilibrium with the tissue compartment partial pressures, with respect to the nonlinear movement of $O_2$ and $CO_2$. Metabolic production of acids, other than carbonic acid via $CO_2$ production, is not modelled. After peripheral tissue equilibration of gases, the venous calculations of partial pressures, concentrations and pH calculations are done using comparable equations as above.

A simple equation of renal compensation for acid base disturbance is incorporated. The base excess (BE) of blood under normal conditions is zero. BE increases by 0.1 per time slice if pH falls below 7.36 (to compensate for acidosis) and decreases by 0.1 per time slice if pH rises above 7.4 (alkalosis).

Each alveolar compartment has a unique and configurable alveolar compliance, alveolar inlet resistance, vascular resistance, extrinsic (interstitial) pressure and threshold opening pressure. For the $i^{th}$ compartment of *N* alveolar compartments, the pressure $p_i$ is determined by:

$$p_i(t_k) = S_i(v_i(t_k) - V_c)^2 + P_{ext,i}v_i(t_k) + P_{INSP} \quad \text{for } i = 1, \ldots, N_A \quad (16)$$

where

$S_i = k_i {N_A}^2/200000$ and $V_c = 0.2V_{FRC}/N_A$

Equation (16) determines the alveolar pressure $p_i$ (as the pressure above atmospheric in $cmH_2O$) for the $i$ th compartment of *N* number of alveolar compartments for the given volume of alveolar compartment, $v_i(t)$ in millilitres. The alveolar compartments are arranged in parallel and interact with the series dead space with respect to the movement of gases. In the case of mechanical ventilation, the flow of air into the alveolar compartments is achieved by a positive pressure provided by the ventilator and the air moves along the pressure gradient. The use of the square of the difference between $v_i$ and *Vc* causes alveolar pressure to increase at volumes below *Vc*, leading to exhalation and a tendency to “snap shut” (mathematical note: the pressure with respect to volume is thus a U-shaped curve) (16).

$P_{ext}$ (per alveolar unit, in $cmH_2O$) represents the *effective net pressure* generated by the sum of the effects of factors outside each alveolus that act to distend that alveolus; positive components include the outward pull of the chest wall, and negative effects include the compressive effect of interstitial fluid in the alveolar wall. Incorporating $P_{ext}$ in the model allows us to replicate the situation of alveolar units that have less structural support or that have interstitial oedema, and thus have a greater tendency to collapse. A negative value of $P_{ext}$ indicates a scenario where there is compression from outside the alveolus causing collapse. The parameter $S_i$ is a scalar that determines the intra-alveolar pressure for a given volume (with respect to a constant collapsing volume $V_c$) and is dependent on the parameter $k$. The units of $S_i$ are $cmH_2O$ $ml^{-2}$. $P_{INSP}$, which represents the pressure generated by the respiratory muscles acting on the lung, is described in the next section. Finally, $V_c$ is defined as a “constant collapsing volume” at which the alveolus tends to empty (through Laplace effects) and represents a fundamental mechanical property of tissue and surfactant (16). $V_{FRC}$ is the end expiratory volume of the lungs.

The effect of the three parameters on the volume–pressure relationship of the alveolar compartments can be observed in Figure S1.2.

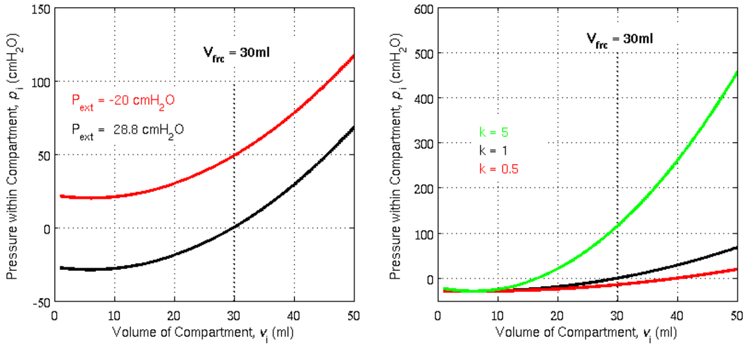

**Figure S1.2 The effect of varying the parameters of Equation (18) on the pressure volume relationship of the model ($P_{INSP}$ = 0) and under mechanical ventilation**

For a healthy lung at the end of the expiration, the ventilator pressure would return to zero above atmospheric (resulting in the tracheal pressure also being equal to zero). The nominal values for ($P_{ext,i}$ , $S_i$ ) have been determined such that at the end of expiration, the alveolar pressure within the compartment is also equal to zero, i.e. at an pre-set value of functional residual capacity.

We consider each of the parameters mentioned above ($P_{ext,i}, S_i$) to be different yet essential components for representing a diseased lung that affect the volume pressure relationship of the alveolar compartments. For example, for a given volume $v_i$ , increasing $S_i$ increases the corresponding alveolar pressure of the alveolar compartment. When compared to another compartment with a lower $S_i$, a larger pressure from the mechanical ventilator would be needed to drive air into the compartment; thus effectively the compartment will be behaving as a stiffer lung unit.

Decreasing $P_{ext,i}$ increases the alveolar pressure such that the pressure gradient (especially during exhaling) forces the air out of the alveolar compartment until the volume of the compartment collapses ($v_i$ = 0 ml). Note that, in effect, the parameters are influencing the resting volume of the compartments (when the alveolar pressure, $p_i$, is equal to zero). If $p_i$< 0 $cmH_2O$, the pressure gradient will cause the flow into the alveolar compartment (as ventilator pressure will always be ≥ 0 $cmH_2O$) until $p_i$ reaches 0 $cmH_2O$.

In the model, the airway resistance $R_{aw}$ is determined by the following equation for N parallel compartments:

$$\frac{1}{R_{aw}} = \frac{1}{R_{B,1}} + \frac{1}{R_{B,2}} + \cdots + \frac{1}{R_{B,N_A}}, \text{for } i = 1, \dots, N_A \quad (17)$$

where $R_{B,i}$ is the bronchial inlet resistance of the $i^{th}$ compartment, which is defined by:

$$R_{B,i} = m_i R_{B0}$$

where $R_{B0}$ corresponds to the default bronchial inlet resistance of an alveolar compartment. $R_{B0}$ is set to $1{\times}10^{-5} \cdot \mathrm{N}$ (the inlet resistance is higher for a model with more compartments as the volume of each compartment decreases) for a healthy lung, giving a resistance of 0.001 kPa per ml per minute for 100 compartments. $m_i$ is a coefficient of the airway resistance, representing a dynamic change in airway resistance and is determined by the equation:

$$m_i = \begin{cases} 1, & \mathrm{t}_{\mathrm{o},i} \leq 0 \\ 10^{10}, & \mathrm{t}_{\mathrm{o},i} > 0 \end{cases} \quad \text{for } i = 1, \ldots, N_A \tag{18}$$

where,

$$\mathrm{t}_{\mathrm{o},i} = \begin{cases} \mathrm{t}_{\mathrm{o},i} - t, & p_{trachea} \geq \mathrm{TOP}_i \\ \tau_{c,i}, & p_{trachea} < \mathrm{TOP}_i \end{cases} \quad \text{for } i = 1, \ldots, N_A \tag{19}$$

$p_{trachea}$ is the pressure in the trachea and $TOP_i$ is a value between 5 and 50 $cmH_2O$ for the $i^{th}$ alveolar compartment. Additionally, a threshold opening pressure (TOP) at low lung volumes needs to be attained for a collapsed alveolar unit to open. Recruitment is a time dependent process, with different airways recruiting at different times, once the threshold pressure has been achieved (17, 18). The equations within the model are solved iteratively as a discretised system. Each iteration represents a physiological time slice of *t* (10 ms). The time-dependent recruitment phenomenon is achieved in the model by the introduction of a parameter $\mathrm{t}_\mathrm{o}$. For collapsed compartments, $\mathrm{t}_\mathrm{o}$ is set to $\tau_c$ which represents the time it could take for collapsed alveoli to open after a threshold pressure is reached. Once $p_{trachea} \geq \mathrm{TOP}_\mathrm{i}$ is satisfied, the counter $\mathrm{t}_\mathrm{o}$decrements during every iteration, and triggers the opening of the airway ($m_i$= 1) as $\mathrm{t}_\mathrm{o} \leq 0$. Otherwise $m_i$ is set to a high value ($10^{10}$) to represent a collapsed airway. We based the range of values for TOP used in these simulations on the work done by Crotti and collaborators (19).

$N_A$ (the number of alveolar compartments) is fixed and set by the user (i.e. they do not change during a simulation). Therefore, during a simulation, $m_i$ , chiefly represents the relatively small changes in inlet resistance during tidal ventilation. Furthermore, $R_{B0}$ are also preset and fixed, and do not change during the simulation. The only change in airway resistance which is dynamic is $m_i$ which is dependent on the volume $v_i$ at time$(\mathrm{t_k})$.

Finally, the pulmonary vascular resistance PVR is determined by

$$\frac{1}{PVR} = \frac{1}{R_{V,1}} + \frac{1}{R_{V,2}} + \cdots + \frac{1}{R_{V,N_A}}, \text{for } i = 1, \ldots, N_A \tag{20}$$

where the resistance for each compartment $R_{V,i}$ is defined as

$$R_{V,i} = \delta_{Vi} R_{V0} \tag{21}$$

$R_{V0}$ is the default vascular resistance for the compartment with a value of $160 \cdot N_A$ dynes s cm$^{-5}$ min$^{-1}$, and $\delta_{Vi}$ is the vascular resistance coefficient, used to implement the effect of hypoxic pulmonary vasoconstriction.

The net effect of these components of the simulation is that the defining, clinical features of ARDS may be observed in the model: alveolar gas-trapping (with intrinsic PEEP), collapse-reopening of alveoli (with gradual reabsorption of trapped gas if re-opening does not occur), limitation of expiratory flow etc.

| **Parameter** | **Size** | **Ranges/Values** | |
|---|---|---|---|
| | | Open alveolar units | Collapsed alveolar units |
| $P_{ext,i}$ | 100 | [-4.8,-6.1] | [2,10] |
| $K_i$ | 100 | [0.4,0.5] | [0.4,0.5] |
| $TOP_i$ ($cmH_2O$) | 100 | [3.5,6.5] | [12,45] |
| Height | 1 | 174 | |
| Weight | 1 | 50+(0.91x(Height–152.4)) | |
| RQ | 1 | 0.85 | |
| I:E | 1 | 1:2 | |
| VO2 (mL.min$^{-1}$) | 1 | 4.2 x Weight | |
| Hb (g.L$^{-1}$) | 1 | 120 | |
| Sh (%) | 1 | 2 | |
| VD (mL) | 1 | 1.75 x Weight | |

**Table S1.1 - List of the parameters required to calibrate the model, with their dimensions and allowable range of variation. $P_{ext,i}$ is the extrinsic pressure acting on compartments; $K_i$ is the stiffness of the compartments; $TOP_i$ is threshold opening pressure of the compartments; RQ is respiratory quotient; VO2 is total oxygen consumption; Hb is haemoglobin; VD is volume of anatomical dead space; Sh is anatomical shunt.**

### Section 2: Modelling spontaneous ventilation

The pressure at the airway opening (e.g. mouth) is relatively constant at atmospheric pressure, while the alveolar pressure value varies during inspiration and expiration. In a healthy individual, the active contraction of the respiratory muscles during inspiration generates a negative alveolar pressure relative to the atmospheric pressure. The variable $P_{INSP}$, which represents the pressure generated by the respiratory muscles acting on the lung, is modelled as a piecewise function as described in (20) and adapted from (21). The function consists of a parabolic profile during the inspiration phase of the respiratory cycle, representing the progressive increase in pressure exerted by the respiratory muscles, followed by an exponential profile during the expiration phase of the respiratory cycle, characterizing the passive relaxation of the muscles. During a single respiratory cycle, $P_{INSP}$ at time $\mathrm{t_k}$, is calculated as

$$P_{INSP}(\mathrm{t_k}) = \begin{cases} \dfrac{-P_{MIN}}{T_I.T_E}.\mathrm{t_k}^2 + \dfrac{P_{MIN}.T}{T_I.T_E}.\mathrm{t_k} & \mathrm{t_k} \in \lfloor 0, T_I \rfloor \\ \dfrac{P_{MIN}}{1 - e^{-\frac{T_E}{\tau}}}.\left(e^{-\frac{((\mathrm{t_k}-T_I)}{\tau}} - e^{-\frac{T_E}{\tau}}\right) & \mathrm{t_k} \in \lfloor T_I, T \rfloor \end{cases}$$

$P_{INSP}$ decreases from zero to its minimum end-inspiratory value ($P_{MIN}$) (i.e., maximum effort) during inspiration and returns to zero at end of expiration. $T$ is calculated from the set respiratory rate, RR, ($T$ = 60/RR). $T_I$ and $T_E$ are the duration of inspiration and expiration, such that ($T$ = $T_I + T_E$). $T_I$ is calculated from ($T_I = T * DC$) where $DC$ is the duty cycle, set to 0.33. $\tau$ is the time constant of the expiratory profile and is set to Te/RR.

The resultant alveolar pressure $p_i$ depends on $P_{INSP}$, as well as on the pressures of the gases within the compartment, the stiffness of the compartment ($S_i$), and parameter $P_{ext}$ which represents extrinsic pressures acting on the compartment.

**Section 3: Modelling Covid-19 Acute Respiratory Failure**

The computational simulator has been configured to represent a number of pathophysiological mechanisms proposed to underlie COVID-19 AHRF. The resulting model replicates the available clinical data (severe hypoxemia combined with preserved pulmonary compliance and gas volumes in 'Type L' ARDS patients). Baseline settings represent a typical 70 kg patient with body mass index of 24 $kg/m^2$.

To model the well-preserved lung compliance seen in type L COVID-19 patients, 8% of the alveolar compartments are modelled as collapsed by increasing the corresponding threshold opening pressures (TOP) and the extrinsic pressures ($P_{ext}$). HPV is normally incorporated in the simulator via a mathematical function, based on the stimulus response curve suggested in (9); to simulate the hypothesised disruption of HPV in CARDS this function has been disabled. Hyper-perfusion was modelled by decreasing the vascular resistance to the collapsed compartments. Pneumonitis is represented by disrupting the gas exchange in 20% of the alveolar compartments. Finally, the effects of microthrombi are implemented by increasing the vascular resistance to perfusion in 10% of the compartments.

### Section 4: Effect of Respiratory effort on compartmental volumes

Figs S4.1 and S4.2 show the effect of increased respiratory effort on the distribution of maximum compartmental volumes. The model consists of 100 independently configured gas-exchanging compartments. Compartments 93 to 100 are collapsed at baseline, at higher respiratory effort some of these compartments are partially recruited. As shown, at the higher values of tidal volume and respiratory rate, the proportion of compartments experiencing higher maximum volumes is significantly increased.

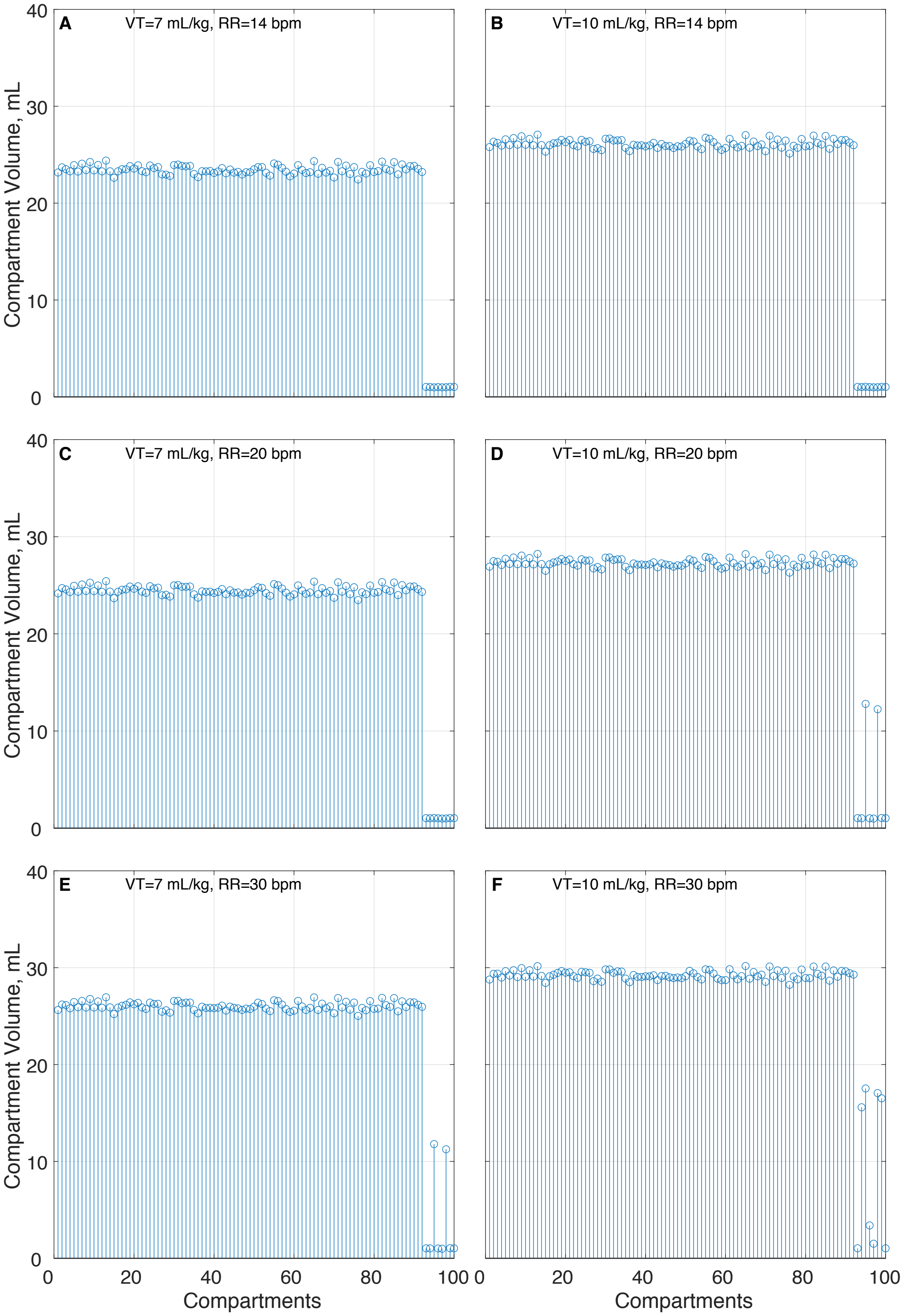

**Figure S4.1 Distribution of maximum compartmental volumes in the model. The model consists of 100 independently configured gas-exchanging compartments. Compartments 93 to 100 are collapsed at baseline, at higher VT some of these compartments are partially recruited.**

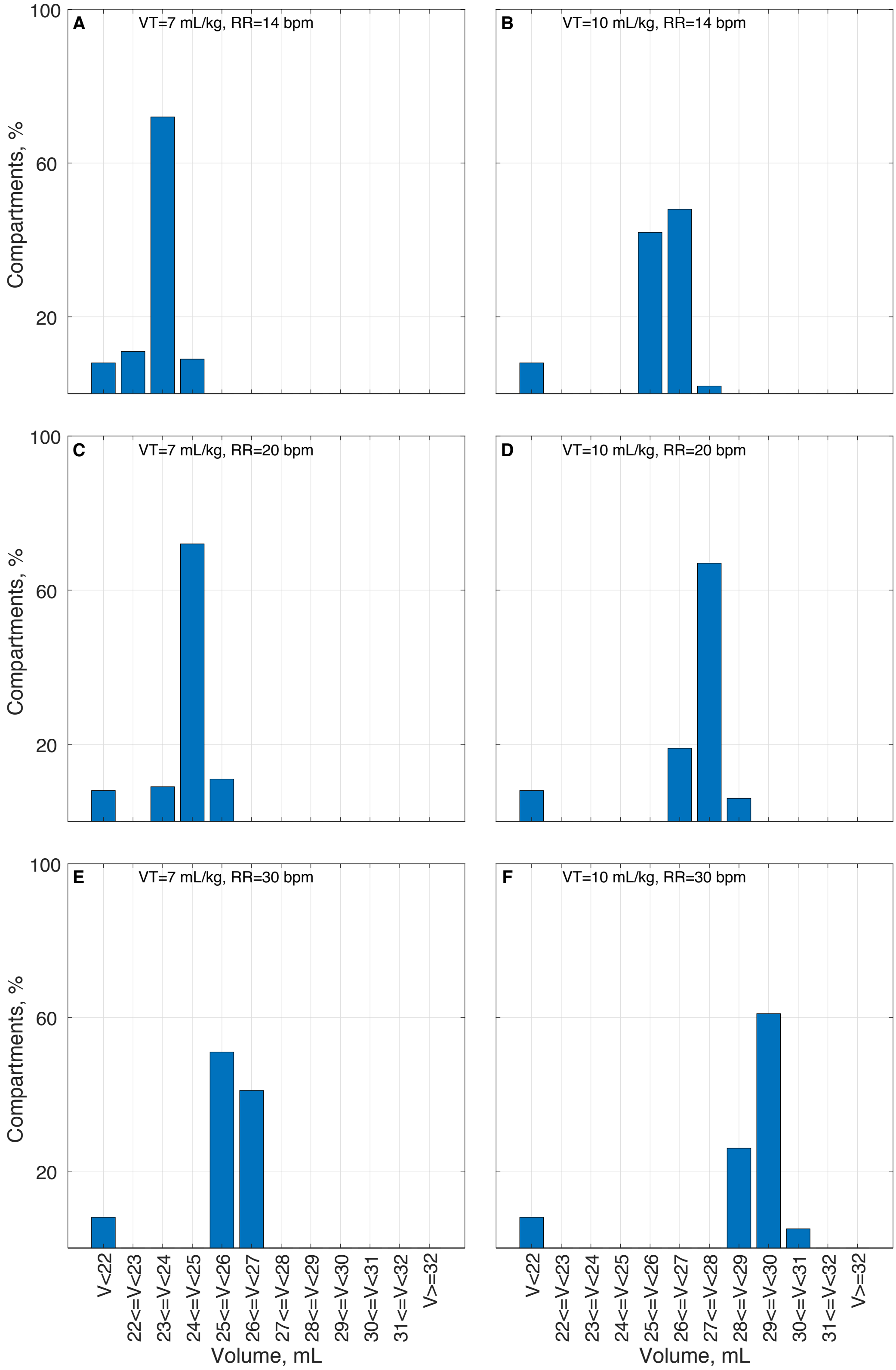

**Figure S4.2: Distribution of maximum compartmental volumes in the model. The model consists of 100 independently configured gas-exchanging compartments. (a) VT = 7 ml/kg, RR = 14 b/min, (b) VT = 10 ml/kg, RR = 14 b/min, (c) VT = 7 ml/kg, RR = 20 b/min, (d) VT = 10 ml/kg, RR = 20 b/min, (e) VT = 7 ml/kg, RR = 30 b/min, (f) VT = 10 ml/kg, RR = 30 b/min.**
